# Voluntary Medical Male Circumcision’s (VMMC) Strategy for HIV prevention in Sub-Saharan Africa, prevalence, risks, costs, benefits and best practice: A scoping review of progress and unfolding insights

**DOI:** 10.1101/2024.06.13.24308912

**Authors:** Charles Maibvise, Takaedza Munangatire, Nector Tomas, Daniel O. Ashipala, Priscilla S. Dlamini

## Abstract

Campaigns to scale up Voluntary Medical Male Circumcision (VMMC) for the prevention of HIV transmission has been going on for years in selected Southern African countries, following recommendations from the World Health Organisations. Despite significant strides made in the initiative and its proven benefits, controversies surrounding the strategy have never ceased, and its future remains uncertain especially as some countries near their initial targets. Over the years, as the campaigns unfolded, a lot of insights have been generated in favour of continuing the VMMC campaigns, while some insights portray the impression that the strategy is not worthy the risks and effort required, or enough has been done, as the targets have been achieved. This article proposes a scoping review that aims at synthesizing and consolidating that evidence into a baseline for a further systematic review aimed at developing sound recommendations for the future of the VMMC strategy for HIV prevention. The scoping review will target all scientific literature published on the Web of Science, Cochrane Library, Scopus, Science Direct, PubMed as well as WHO Institutional Repository for Information Sharing (IRIS) since 2011. The review shall be guided by Arksey and O’Malley’s (2005) framework for scoping reviews, and the Preferred Reporting Items for Systematic Reviews and Meta-Analyses Extension for Scoping Reviews (PRISMA-ScR) checklist shall be followed. Discussion of the findings is envisioned to yield evidence that can be further analysed to give insights about risk/cost-benefits ratios of the strategy at this point in time as well best clinical practices for the VMMC procedure, to inform the future of the strategy.

## INTRODUCTION

Over that past two decades, Voluntary Medical Male Circumcision has been among the leading interventions in HIV prevention globally, as a complement to the conventional strategies. This followed a recommendation by the World Health Organization that VMMC be added to the comprehensive package for HIV prevention (1). In particular, the Eastern and Southern African Countries have been the core of VMMC activities, in view of the then relatively low VMMC prevalence coupled to high HIV prevalence. In this regard, a total of 15 countries embarked on massive VMMC scaling-up. Initially, these were Botswana, Eswatini, Ethiopia, Kenya, Lesotho, Malawi, Mozambique, Namibia, Rwanda, South Africa, Uganda, Tanzania, Zambia, and Zimbabwe (2). Later on, the Gambela province of Ethiopia was added (3), and of late, South Sudan (4).

The World Health Organization has been continuously issuing guidelines and practice manuals which countries have been adapting and adopting to their local context. Figure 1 highlights some of the main reference documents issued by the Organizations and the key message or update in each of them (1,5–11).

**Fig 1.**
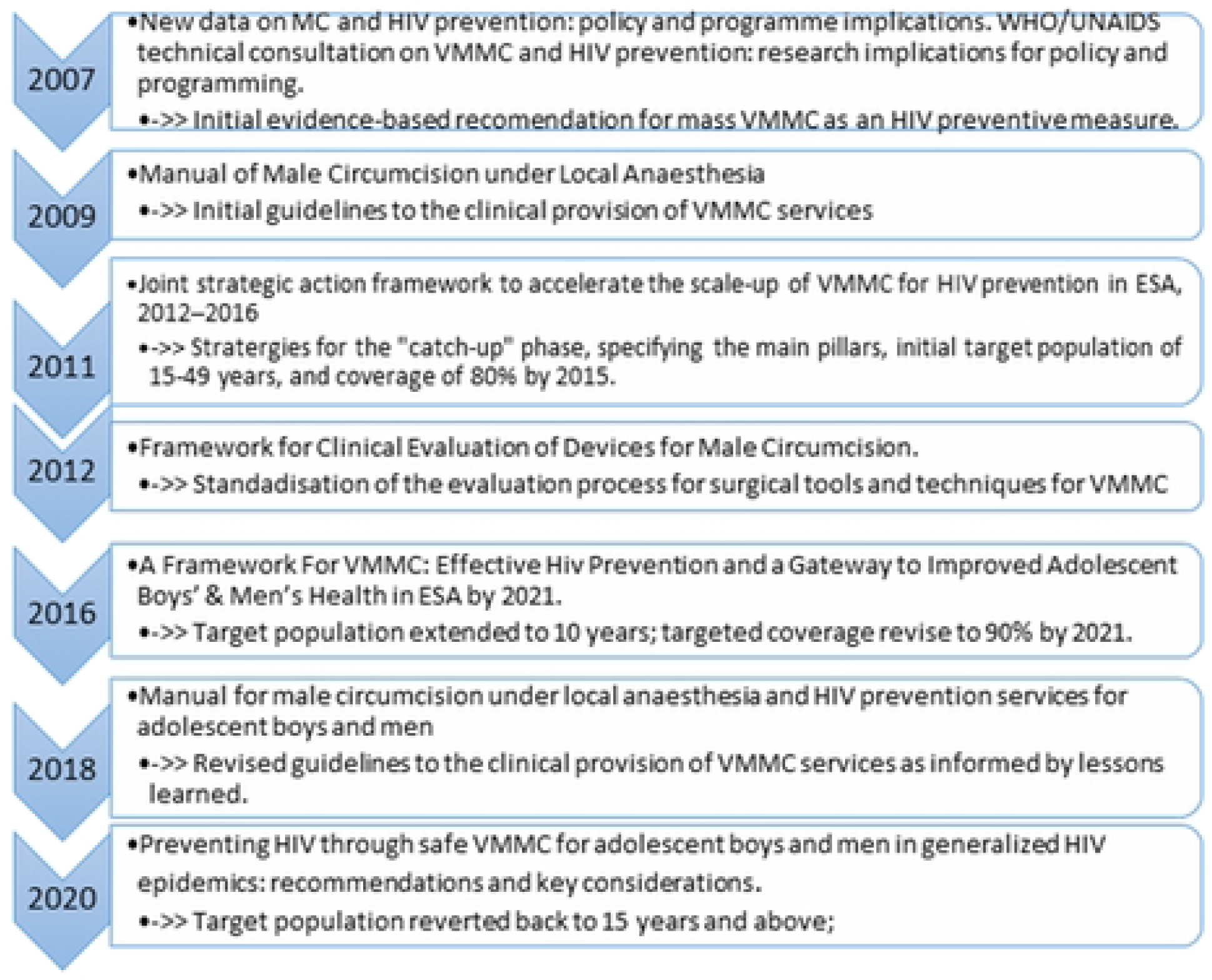
Sequential Guiding documents for VMMC

From its inception, VMMC campaigns have been marked with multiple knowledge gaps and misconceptions, rendering it a significantly controversial strategy, thereby prompting multidimensional empirical enquiries. Some of the key focal grey areas included its impact on sexual performance, pleasure and satisfaction thereof (12,13); risk compensatory behavior (14–16), as well as prevalence and severity of adverse events (13,17). To date, successes, challenges and future projections in the VMMC campaigns are essentially a function of evolution of insights regarding the aforementioned grey areas. Noting also that the VMMC strategy is beneficial only in high HIV epidemic areas with low MC prevalence (18,19), it follows that hypothetically the cost-benefit ratio dwindles with time as set targets are met, unless otherwise influenced by unfolding insights.

The primary target of VMMC campaigns was to pull up the global prevalence of male circumcision from around 30% in 2007 (20) to about 80% by 2015, particularly among men aged 15 to 49 years in the targeted ESA countries, in the so called “catch-up” phase (5). This target was later revised to 90% by 2021 (7). Notably these targets were not met, for various reasons, and a decrease in momentum has been recorded, though hopes still remain (4). The circumstances surrounding these undesirable outcomes are not clear. Whatever the cause, it adds onto the uncertainties of the future of VMMC campaigns. By default, it could be attributed to perceptions a decrease in relative epidemiological benefits as the initiative nears its targets, that is, an increase in the prevalence of male circumcision and a decrease in the burden of HIV globally. However, it is worth noting that data suggests that there are other emerging benefits of VMMC, other than HIV prevention, that were not considered in the initial modelling of the cost/risk-benefit ratio of the strategy (2,19,21). Likewise, there are also some emerging complications and/or adverse effects that are being realized (22,23). A closer analysis and synthesis of these insights is therefore necessary to inform future dimensions in VMMC, and hence this proposed scoping review.

Scoping reviews have gained significant momentum as a rapidly emerging method for synthesizing evidence across diverse domains (24,25). Thus, this article intends to explore updates relating to the VMMC strategy for HIV prevention in order to establish a clear picture of the status quo. Insights generated from the field and the subsequent policy recommendations from time to time as well as progress made to date would be explored with a view to pooling evidence worth considering in determining the future of the VMMC strategy. Overall, this evidence is meant to form the basis for a systematic review study aimed at projecting and recommending sound prospects for the strategy.

Specific research objectives for this study are:

- Assess the prevalence of male circumcision in Sub-Sahara Africa?
- Explore and synthesize evidence on VMMC programmes in terms of risks, costs, benefits and best practices.

## SIGNIFICANCE

The study will consolidate available data and give a comprehensive impression of VMMC activities to date, starting with progress made so far. Emerging and unforeseeable insights as well as answers to historical grey areas regarding selected risks, costs and benefits will be compiled as well. Thus, other potential benefits of VMMC complementing the HIV prevention role will be unveiled. Similarly unforeseeable complications, adverse effects, risks and/or costs will also be revealed. This will form the basis for a more comprehensive and updated cost-benefit analysis in order to determine the soundest course of action for the VMMC strategy going forward. Programmatically this will foster effective utilization of available resources, thus investing in VMMC only if the benefits are worth the effort. Recommendations based on the study will also ensure that, in the continuation of VMMC, best practices are adopted in the interest of safety and cost effectiveness.

## MATERIALS AND METHODS: SCOPING REVIEW

This protocol is for a scoping review of literature reporting on progress and updates on the VMMC strategy for HIV prevention, as well as insights generated from its implementation. Scoping review method is suitable since its goal is to synthesize different types of evidence on a particular area and identify gaps for future research (26). Overall, the Preferred Reporting Items for Systematic Reviews and Meta-Analyses Extension for Scoping Reviews (PRISMA-ScR) checklist (27) will be used in this review. The methodology will be guided by the Arksey and O’Malley framework for scooping reviews (28), and the steps to be followed are identifying the research question, identifying relevant studies, selection of eligible studies, recording the data, chatting and summarising findings and voluntarily expert consultation (Figure 2).

**Fig 2.**
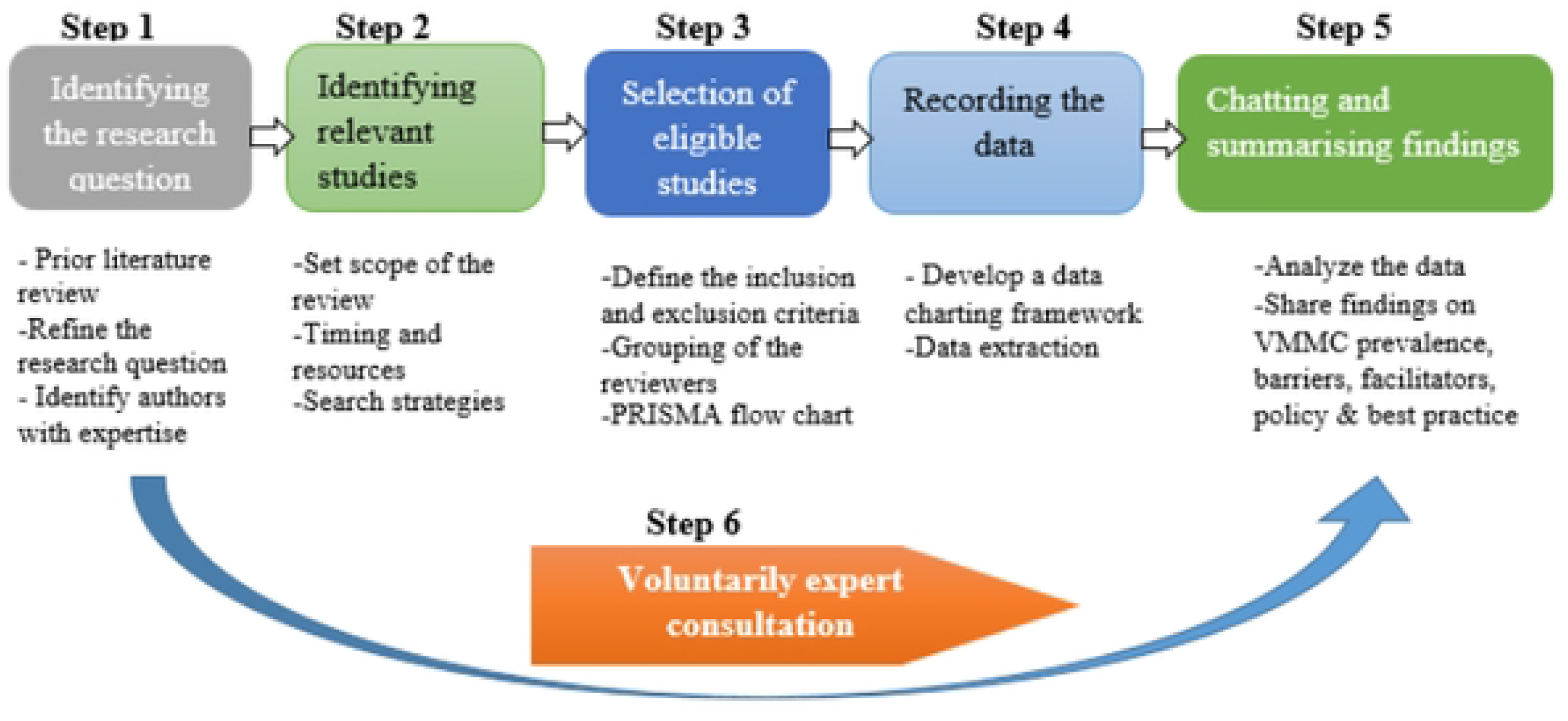
Visual representation of Arksey and O’Malley framework

### Step 1: Identification of the research question

The “PCC” framework for developing a research question was used in this study (29,30). The population (P) of interest are men and children aged ten (10) years or older, being the primary targets of VMMC at some point. The concept (C) under study is the VMMC strategy for HIV prevention, with a particular interest in gained insights that are worth considering in determining the future of VMMC. The Context (C) for the study is Sub-Saharan Africa, where the strategy was rolled out. The overall research question for this review is “What insights have been gained regarding risks, costs and benefits of VMMC as a strategy for HIV prevention in Sub-Sahara Africa?” The aim is to ascertain progress made so far and to pool the evidence worth considering in determining the future of the VMMC strategy for HIV prevention. The specific research questions, therefore, are as follows:

- What is the prevalence of male circumcision in Sub-Sahara Africa?
- What evidence has been generated regarding VMMC in terms of risks, costs, benefit and best practice in the performance of the procedure?

### Step 2: Identifying relevant studies

A comprehensive literature search will be conducted, utilizing selected electronic databases, namely: Web of Science, Cochrane Library, Scopus, Science Direct, PubMed as well as WHO Institutional Repository for Information Sharing (IRIS). These sources were selected based on the relevance of their scope to the concept under investigation. Initially, key words that shall be used, singly and in combination, are: “Circumcision”, “VMMC” and “Sub-Sahara Africa”. Only studies published in English between 2011 and 2024 will be considered, based on the fact that the VMMC strategy was rolled out into full force around 2011 in most countries (31). To ensure the effectiveness of our search strategy, a pilot study will be conducted to assess the relevance of the keywords and databases in addressing our research inquiries. Any necessary adjustments will be made accordingly.

### Step 3: Selection of eligible studies

Two researchers will conduct a rigorous assessment of citation titles and abstracts, as well as thoroughly examine potentially pertinent articles, utilizing the inclusion and exclusion criteria stipulated in table 1. The criteria were framed according to the PCC framework. The authors will exclusively focus on articles that present original research on Voluntary Medical Male Circumcision (VMMC) in Sub-Sahara Africa. In the event that there is a disagreement between the reviewers after reviewing the abstract or full article, a third reviewer will be consulted to provide an expert opinion.

**Table 1:** Inclusion and exclusion Criteria.

|  | **Inclusion Criteria** | **Exclusion Criteria** |
| --- | --- | --- |
| **Population** | Rereview targets literature reporting on men and children aged 10 years and older since these were the targets for VMMC programme. | Studies focusing on other age groups will be excluded. |
| **Concept** | Literature on VMMC will be included, particularly that which gives insights worth considering in determining the future of VMMC. These include “Complications”, “Risks”, “Benefits”, “Costs”, “Recommendations” and “Coverage”, or Prevalence”, that is, “Progress towards set targets.” | Literature that gives no insights that can form the basis of arguments for or against VMMC, or progress towards set targets, will be excluded |
| **Context** | Only literature on circumcisions performed in Sub-Saharan Africa under the VMMC programme will be included. The “voluntary” and “medical” aspects are key | Literature on traditional or religious male circumcision and Early Infant Male Circumcision will be excluded. These contexts have a different profile of risks, benefits, ethics, and other variables that are significant in VMMC |
| **Type of sources** | All literature published in the targeted databases.<br>All types of original research and review articles will be included.<br>Quantitative, qualitative and/or mixed-method study design will be included;<br>Only papers written in English and published between the period of 2011 – 2024 | Literature published in platforms other than the targeted databases will be excluded.<br>Media reports and personal views will be excluded |

The Joanna Briggs Institute’s (JBI) PRISMA flow diagram for scoping reviews will be followed, as illustrated in figure 3 (30). Once retrieved the articles will be entered into Mendeley Reference Manager and duplicates will be eliminated. Then the screening of the articles using title and abstracts will be done to determine eligibility of the studies for this review. In cases where full text is not readily available librarians and authors will be reached for assistance. All the authors will be involved in the full text screening and any disagreements will be reached through a consensus process and not majority vote and this process will be documented.

**Fig 3.**
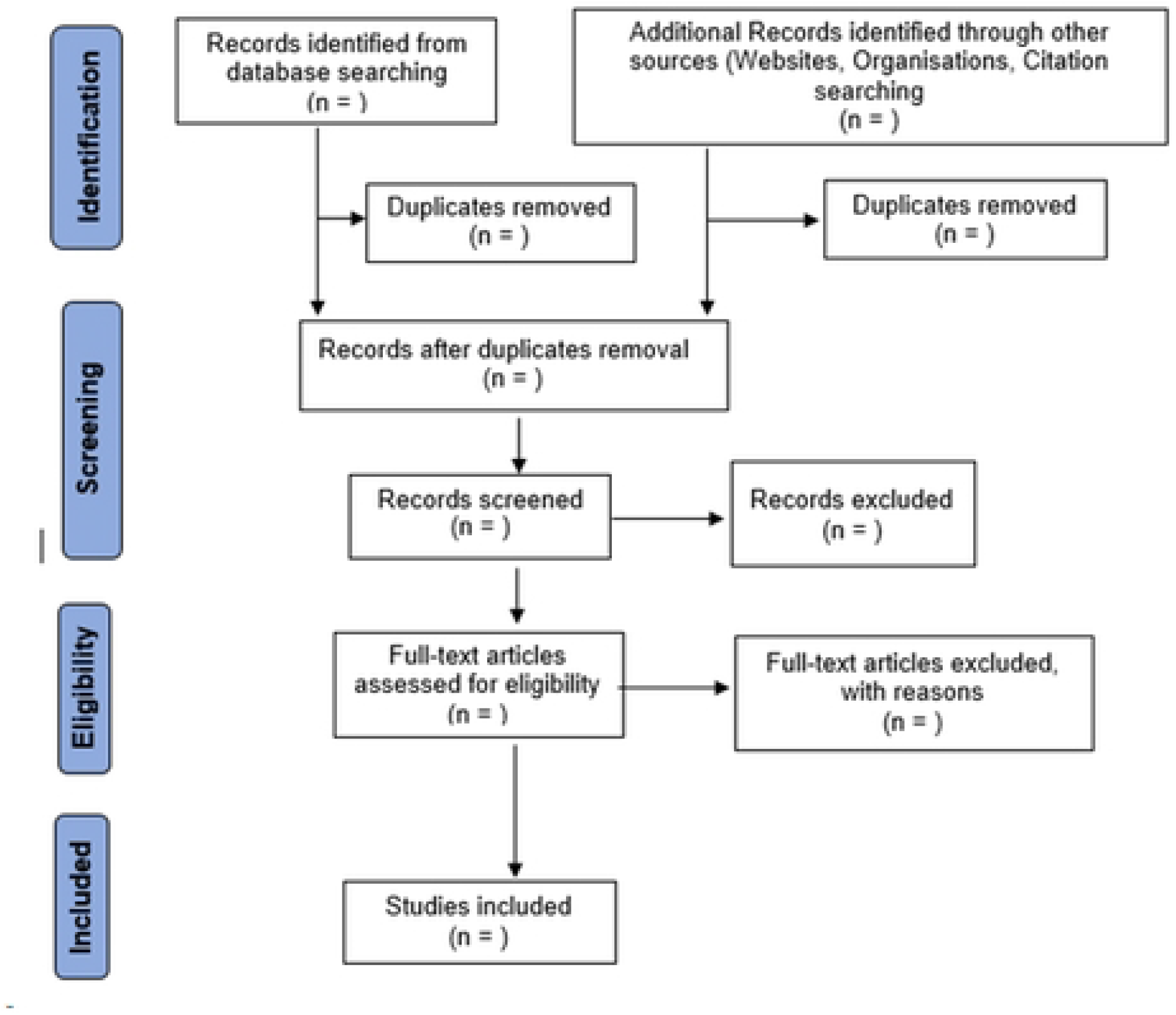
PRISMA flow diagram for scoping reviews [Adapted from JBI, (2015) (30)]

### Step 4: Charting the data

Data will be extracted from full-text articles using the data extraction template presented in table 2. Variable in the template were adapted from the JBI (2015) reviewer’s manual for scoping reviews and complemented by the work of Widyaningsih et al (32). The template may be revised per rising need as informed by the data collection process. Three authors will independently extract the data onto the electronic version of the form. Though the inclusion of quality appraisal in scoping reviews remains debatable (33), a formal critical appraisal of primary studies will be done since findings of the review may potentially influence practice (30). The Mixed Method Appraisal Tool (MMAT) for scoping review shall be used for that purpose (34).

**Table 2:** Data extraction template (Adapted from JBI, 2015)

| **Data** | **Data description** |
| --- | --- |
| Details of the article | Author (s), year of publication, location of the study |
| Title | Full title of the article |
| Type of source | Peer reviewed journal, grey literature, international guidelines, report |
| Study design/method | Quantitative; Qualitative; Mixed method |
| Aim/purpose | Overall aim or objective of the study |
| Population and sample size | The population targeted in the study and the sample size if applicable |
| Type of intervention | Details of intervention and/or comparator if applicable. |
| Concept of VMMC covered or reported | Relevant data extracted from the source, such as VMMC prevalence; risks; costs; complication; benefits; recommendations for best practice. These concepts shall constitute major categories or themes of the findings. |
| Results/findings | Each concept reported in the results/findings shall be analysed and its overall impression and implication for VMMC shall be presented and classified under the respective category as a sub-theme. |
| Additional information | Any additional information worthy considering, e.g. limitations of the study, validity and trustworthiness of findings/ |

### Step 5: Collating, summarising and reporting the results

The PRISMA flow diagram (Fig 1) shall be used to summarise the review process, and a summary table of all included studies shall be formulated, capturing the variables outlined in the data extraction template (Table 2). The charted data will be synthesized and thematically analysed, and a narrative report of the findings compiled. The PRISMA-ScR checklist will be used to guide the writing of the overall report for the review (27).

### Step 6: Conducting consultation

While it is acknowledged that this step may be optional in a scoping review (28,30), there is growing evidence affirming the benefits of this step (33). As such, consultations with experts and key stakeholders in the VMMC campaigns shall be conducted. Findings of the review shall determine the most appropriate informants in this step, but tentatively, custodians of the VMMC campaigns at national and regional levels shall be targeted. The purpose of the consultations will be to validate the findings as well as soliciting additional insights that may have been missed from the primary data sources consulted (33).

### Ethical consideration

There is no ethical approval require since data will be collection from reviewed literatures as opposed to individuals

### Registration of protocol

This protocol is registered with the Open Science Framework, registration ID https://doi.org/10.17605/OSF.IO/SFZC9.

## DISCUSSION

This scoping review focus on mapping literature on voluntary male circumcision is Sub-Saharan Africa with the goal of synthesizing the evidence around risks, benefits and best practices. It is hoped that the review will consolidate cumulatively available evidence on progress, risks, costs and/or benefits of VMMC thereby setting a stage for further analysis aimed at informing the direction of future policy, research and practice regarding. The future of the VMMC strategy lies mainly on goal attainment, that is, VMMC coverage relative to the set targets, perceived risk-benefit ratio at individual level as well as presumed cost-benefit ratio programmatically at national and regional levels. Overall, these factors determine the worth and future prospects of the strategy. To date, there are still numerous dilemmas, mixed feelings, knowledge gaps as well as underutilized emerging insights and discoveries surrounding these aspects of the VMMC strategy (13,21). It is envisioned that this review will bring to fore the much-needed insights and consciousness regarding those aspects, and hence form the basis for further studies to develop policy recommendations relating to the future practice of VMMC for HIV prevention in the region.

## Data Availability

Deidentified research data will be made publicly available when the study is completed and published.

doi:10.7326/M18-0850

## LIMITATIONS

In this review, only published literature available online in selected databases and in reputable organizations and Government Departments will be considered. Thus, potentially valid evidence outside these sources is excluded from the synthesis. However, choice of the sources followed careful and purposive consideration to maximize inclusivity. As with most scoping reviews, the quality of the evidence accessed will not be assessed hence the results will have to be interpreted with this understanding. In any case, the chosen sources are known for credibility.

## SUPPORTING INFORMATION

S1 – PRISMA ScR Checklist

## AUTHOR CONTRIBUTIONS

**Conceptualization:** C. Maibvise, T. Munangatire

**Development of protocol:** C. Maibvise, T. Munangatire, N. Tomas, D.O. Ashipala, P.S. Dlamini

**Project administration:** C. Maibvise, T. Munangatire

**Data extraction:** N.Tomas, D.O. Ashipala, P.S. Dlamini

**Data analysis:** C. Maibvise, T. Munangatire, N.Tomas, D.O. Ashipala, P.S. Dlamini

**Report writing** – Initial draft: C. Maibvise, T. Munangatire,

**Finalising report**. C. Maibvise, T. Munangatire, N.Tomas, D.O. Ashipala, P.S. Dlamini

## Notes

### Competing Interest Statement

The authors have declared no competing interest.

### Funding Statement

The author(s) received no specific funding for this work.

### Author Declarations

None / Not applicable

